# RAPEX HARM from AIS 2015 Coded Injuries

**DOI:** 10.64898/2026.03.04.26346267

**Authors:** J. Krampe, M. Junge

**Affiliations:** TU Braunschweig, Germany; Volkswagen AG, Wolfsburg, Germany

**Keywords:** RAPEX, HARM, AIS 2015, injury aggregation, long-term consequences

## Abstract

The European Union’s Safety Gate Rapid Alert System (RAPEX) requires Hazard and Risk Assessment Methodology (HARM) evaluations addressing both injury lethality and long-term consequences (LTC). This paper developed a post-processing method to use AIS 2015-coded trauma data directly for RAPEX HARM assessments.

AIS 2015 utilizes two metrics: the AIS Code (AIS-CD) for threat to life and the predicted Functional Capacity Index (pFCI) for LTC. While the AIS-CD has been validated on numerous datasets, the pFCI values are based on a theoretical framework that is pending validation.

To counter coding variability and poor alignment with clinical diagnoses, initial AIS identifiers (AIS-IDs) were aggregated to a robust level of detail for both metrics. Individual injury severities (AIS-CD/FCI-CD) were aggregated to the person level using a conversion derived from the three most severe injuries (triples), mirroring the concept of the New Injury Severity Score (NISS). The final HARM Level is the most severe outcome derived independently from either the AIS-CD or FCI-CD triple conversion.

Analysis showed over 70% of injuries in the aggregated codebook had no LTC. While AIS-CD dominated lower HARM scores, LTC became more defining with increasing HARM severity for the GIDAS sample. At HARM 4 (highest severity), AIS-CD accounted for 53% of cases, FCI-CD accounted for 16%, and both were equally severe in 31% of cases. This method successfully assigns HARM values to AIS 2015 injuries, providing a more holistic severity measure than the current AIS-CD-only approach.

**Highlights:**

- Novel method assigns RAPEX HARM values to AIS 2015-coded injuries.
- Combines lethality (AIS-code) and long-term consequences (FCI-code) for injury severity assessment.
- Aggregates injury severity using the three most severe injuries per person.
- Long-term consequences account for 13% and 16% of the highest two HARM ratings, respectively.

## 1. Introduction

The Rapid Exchange of Information System (RAPEX) describes a method for detecting unsafe consumer products.[1] With its introduction, the European Union emphasized the protection of consumers from health hazards and technical defects in defective products.

We developed a post-processing method for AIS 2015-coded[2] trauma data, such as the DGU Trauma Register[3], GIDAS[4], or NASS CISS[5], that combines the injury lethality and long-term consequences of individually coded injuries and aggregates this information to the patient level for direct evaluation in RAPEX HARM assessments. Thus, this provides the ability to use AIS 2015-coded injury data for RAPEX evaluations, i.e., clustering accident and crash data into risk-specific groups, such as speeding, rear-end collisions, or mobile phone use.

The method is demonstrated on real-world crash data from the GIDAS[4] database.

## 2. Material

### 2.1. AIS 2015 Codebook

The 2015 edition of the Abbreviated Injury Scale (AIS) was the basis of this investigation.[2] It provides the total enumeration of codeable individual injuries via the unique identifiers (AIS-IDs).

To better align the AIS-IDs with medical coding by clinicians, some very specific AIS-IDs were reduced to an unspecific ‘not further specified’ (NFS) AIS-ID already in the AIS codebook. Some newly introduced AIS-IDs are just clusters of AIS-IDs already in the codebook.

An AIS Code (AIS-CD) and a ‘predicted functional capacity index’ (pFCI) coding for the lethality and the long-term impairment, respectively, are attached to each injury in the AIS 2015 codebook.

### 2.2. GIDAS Database

Without loss of generality, the AIS 2015 coded medical data from the German in-depth Accident Study (GIDAS)[4] were used as a testbed for calculating the HARM values.

The current GIDAS dataset aims to be a 50% sample of real-world crash situations involving at least one injured participant in the Greater Dresden and Greater Hannover areas. The sample size is about 1,000 crashes per metropol-region per year. We used crashes from 1999 to 2024 for our evaluation.

The medical information was used independently of the type of crash involvement, i.e., whether the individual was a vehicle occupant or a vulnerable road user.

## 3. Methodology

The fundamental idea behind AIS coding of injuries is to segregate a patient’s injuries into individual entities, known as AIS-IDs, and assess their severity separately. The segregation process is detailed in the AIS codebook.[2] Two individual injury severity assessment schemes are currently detailed in the AIS codebook: While the Abbreviated Injury Scale, i.e., the AIS-CD, focuses primarily on the threat to life, the pFCI measures long-term impairment if the injury is survived.

Individual injury assessments must be aggregated to evaluate injury severity at the patient level.

Injuries from traffic crashes are primarily the result of blunt force trauma.[6] Therefore, this evaluation focuses on said injuries, deemphasising penetrating and blast-related injuries.

### 3.1. AIS-ID Aggregation

The AIS 2015 codebook distinguishes between 2,000 individual injuries, each assigned a unique 6-digit numerical identifier (AIS-ID).[2] The codebook represents an evolutionary advancement of the AIS 90[7], AIS 90 Update 98[8], and AIS 2008[9] codebooks. Its focus shifts from lethality to include injury-related functional incapacitation, thereby increasing the number of AIS-IDs from 1,300 to over 2,000. Although the codebooks provide comprehensive coding rules, the AIS-IDs do not align well with medical diagnoses and treatment regimens, i.e., the coding of injuries used by medical doctors and other professionals involved in the care of trauma patients.[10]

The AIS 2015 codebook treats the AIS-ID as a key variable that codes for individual injuries, to which an AIS code (AIS-CD) and a pFCI value are attached. While the AIS-CD represents the injury’s lethality, the pFCI codes the long-term incapacitation resulting from the injury if the patient survives.

#### 3.1.1. AIS-Code

The AIS-CDs range from 1 (minor) to 6 (maximal), and 9 for ‘unknown’.[2] Uninjured body regions are usually coded as 0 (uninjured).

The AIS-CDs from 0 to 6 are highly correlated with lethality.[11]

#### 3.1.2. AIS-ID Aggregation for the AIS-Code

In a recent study, we aggregated AIS-IDs to address misalignment between AIS-ID coding and medical diagnoses coding,[10] as well as the tendency of coding personnel to up-or downcode, i.e., assigning AIS-IDs with a higher or lower AIS-CD depending on their impression of the case’s severity[12].[11] One reason for the misalignment mentioned above is the coding of AIS-IDs from the provided medical documentation, which means no further diagnostic tests can be ordered, and no further questions can be asked. We introduced three levels of detail, with Level III being differentiated from Level II on morphological grounds. At Level III, the coder can differentiate between contusion, laceration, avulsion, and thrombosis, and distinguish between brain swelling and edema; they can also diagnose a traumatic aneurysm, etc.

The distinction between Levels I and II focuses on diagnostics. At Level II, the coder can distinguish between a subluxation and a dislocation, a herniation and a rupture, and recognize transient, incomplete, and complete neurological signs. Additionally, they can identify named arteries, veins, and cranial nerves. Level I coding stops at the organ system level, requiring no further differentiation; it is coded as ‘not further specified’ (NFS).

The effect of this AIS-ID aggregation on AIS-CD distribution is shown in Table 3.

##### AIS 1 Skin Injuries

Most AIS 1 skin injuries are self-reported and not documented in medical records, as they are not reimbursable. Relying on self-reported injuries results in bias, i.e., overreporting by those who think they can claim compensation for their pain and suffering. Furthermore, AIS 1 skin injuries dominate other AIS 1 injuries, especially those in the distal parts of the extremities. Thus, we elected to scrub AIS 1 skin injuries from the evaluation, including AIS 1 penetrating injuries.

Note that even in the AIS codebook, all skin injuries have an FCI-Code of 1. At Level 1, all skin and penetration injuries have an AIS-Code 1 and an FCI-Code of 1, i.e., ‘perfect state’ (see below), assigned. Thus, these injuries would not change the HARM rating (see Table 2). In Figure 1, the influence of scrubbing skin injuries on the S-scale can be seen in the leftmost bars for the lower S-scale values.

**Figure 1.**
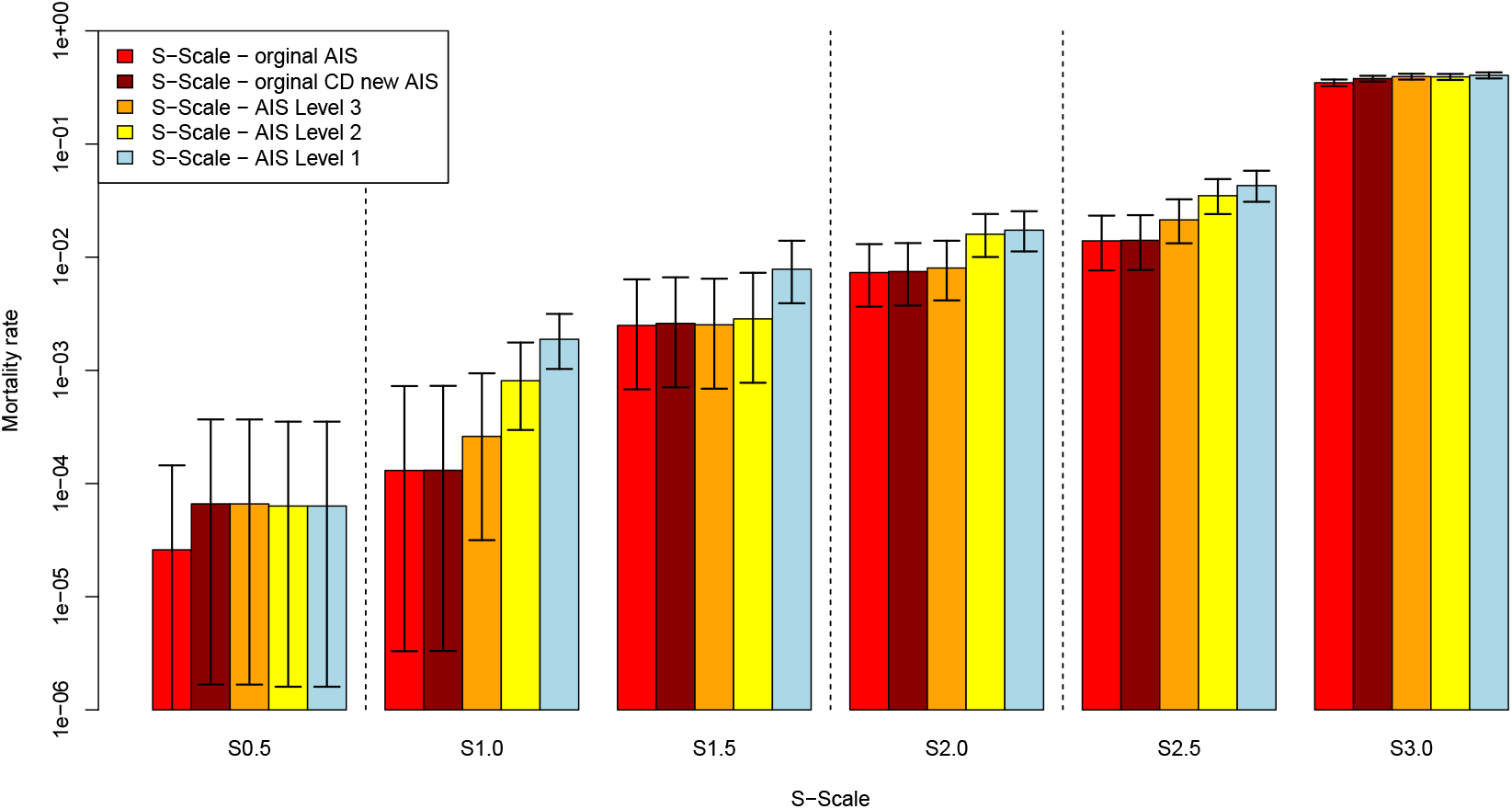
Mortality of the S-scale at different AIS-ID aggregation levels for the GIDAS dataset. The S-scale mortality is statistically significantly different at Level 1 (light blue, rightmost columns). The increase in mortality at the S 0.5 level from the ‘original AIS’ is due to the exclusion of superficial skin injuries; the small increase from the ‘original AIS’ bar at S 3.0 results from the AIS-CD 9 processing. The increase in mortality with increasing AIS-ID aggregation can be seen as a reversal of upcoding: the coarser the injury description, the lower the AIS code. The dotted vertical lines show the clustering for the AIS-CD based HARM values.

#### 3.1.3. FCI-Code (FCI-CD)

For every individual injury in the AIS 2015 codebook, i.e., every AIS 2015-ID, a *‘predicted Functional Capacity Index’* (pFCI) value coding the functional limitation following non-fatal injuries is given. The pFCI was determined by an iteratively by a panel of experts[13] for the AIS 98[8] and AIS 2008[9] codebooks and subsequently extended to the AIS 2015 codebook.[2]

We recoded the pFCI to the *‘Functional Capacity Index’* code (FCI-CD) for ease of use and better alignment with the AIS-CD metric so that 1 corresponds to the ‘perfect state’ and 5 to the ‘worst possible state’:

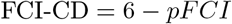

The FCI-CD assesses the *long-term consequences* (LTC) of an individual injury.

#### 3.1.4. AIS-ID Aggregation for the FCI-Code

We developed an AIS-ID aggregation for the FCI evaluation independent of the AIS-ID aggregation for the AIS-CDs.

The focus was on easily diagnosable loss of functionality, such as motor loss, deafness, and blindness, as well as on injuries with potential future impairment, including joint and bone injuries with cartilage involvement. We refer to this aggregation as the AIS-ID Level 1 FCI aggregation. This aggregation reduces the number of AIS-IDs from over 2,000 to 453.

Note that the AIS-ID aggregation for AIS-CD and FCI have different foci. While the AIS-ID aggregation for AIS-CD primarily concerns the acute lethality of an injury, the aggregation for the FCI focuses on the long-term consequences (LTC) of the injury, assuming the patient survives the trauma. While AIS-IDs relevant to AIS-CDs can be extracted from emergency medical protocols and hospital discharge letters, functional deficiencies are best coded from physical examination records.[14] At the same time, cartilage damage is well documented in radiological reports.

### 3.2. Person Level Aggregation

Historically, the AIS-CD was aggregated to the patient level by simply taking the maximum AIS-CD, i.e., MAIS, as the patient’s overall injury severity.

Even though the Injury Severity Score (ISS)[15], which specifies six body regions and combines the three most severely injured regions into a single ‘score’, highlighted problems with the MAIS, its complicated calculation and susceptibility to coding errors hindered its broad adoption. As the AIS-CD (and also the FCI-CD) are ordinal-scale metrics, a scale transformation to a metric or interval scale is necessary to perform arithmetic operations in a mathematically sensible way. While the ISS definition used the simplest non-linear function, squaring, for this transformation, we showed in previous papers that an exponential transformation provides a more linear correlation with mortality.[16][11]

To elaborate, let *AIS*_*i*_ be the AIS-CD of injury *i* = 1, …, *n*, where *n* refers to the *n* injuries a person sustained. Then, the AISx is defined as

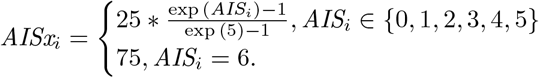

Additionally, let *AISx*_[*i*]_ be in decreasing order such that *AISx*_[1]_ refers to the most severe injury. Using this transformation and focusing, as in the New Injury Severity Score (NISS),[17] only on the three most severe injuries of a patient, we derive

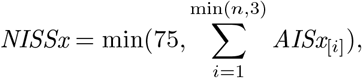

where the outer minimum keeps the NISSx in the range of 0 to 75.

Note that the NISSx scale is monotonous with injury lethality and that the transformation is bijective, i.e., there is precisely one AIS-CD-triple that matches one NISSx value, as opposed to the NISS definition. Thus, a direct mapping of AIS triples to injury severity levels (S-levels) can be derived, as shown in Table 1.[11] The NISSx cut-off values were defined so as not to separate 0 and 1 codes, except for the S0 — S0.5 division. For the HARM-levels, the S-levels were aggregated further, see Table 2.

**Table 1:** Definition of the S-Scale.[11] Sorted triples, i.e., *x* is less or equal to the severity to its left.

| S-Level | NISSx | code triple | description |
| --- | --- | --- | --- |
| 0.0 | $\geq 0.0$ | 0/0/0 | ‘no’ (significant) injuries |
| 0.5 | $> 0.2$ | 1/$x$/$x$ | minor injuries |
| 1.0 | $> 1.0$ | 2/0/0, 2/1/$x$ | light injuries |
| 1.5 | $> 2.0$ | 2/2/0, 2/2/1 | moderate injuries |
| 2.0 | $> 3.0$ | 2/2/2, 3/0/0, 3/1/$x$ | severe injuries |
| 2.5 | $> 4.0$ | 3/2/$x$ | life-threatening injuries (survival probable) |
| 3.0 | $> 5.5$ | 3/3/$x$, 4+/$x$/$x$ | life-threatening injuries (survival uncertain) |

**Table 2:** Code-triple to HARM conversion for sorted AIS-CD and FCI-CD triples, i.e., *x* is less or equal to the severity to its left.

| HARM | S-Scale | AIS-CD and FCI-CD Triple |
| --- | --- | --- |
| 1 | S 0.0, S 0.5 | 0/0/0, 1/$x$/$x$ |
| 2 | S 1.0, S 1.5 | 2/0/0, 2/1/$x$, 2/2/0, 2/2/1 |
| 3 | S 2.0 | 2/2/2, 3/0/0, 3/1/$x$ |
| 4 | S 2.5, S 3.0 | 3/2/$x$, 3/3/$x$, 4+/$x$/$x$ |

Note that we use sorted triples throughout, i.e., the highest code in the triple is to the left.

#### 3.2.1. AIS

##### AIS-Code Aggregation

Patients with AIS-coded injury severities ranging from 0 to 6 were aggregated to the patient level using Table 2, i.e., by considering the three most severe injuries as representative of the patient’s overall injury severity. Cases with an AIS 9-coded injury received special treatment (see below).

##### AIS 9 Code: minimum AIS

Some AIS-IDs code for injuries that have not been fully diagnosed, e.g., *vascular injury to the face*. Although a more detailed diagnosis would be possible, it has not been done or documented. The AIS-codebook assigns an AIS-code 9 (‘unknown severity’) to these AIS-IDs.

For each AIS-ID, the minimum AIS (mAIS) code needed to inflict such injury was determined. The mAIS ranges from AIS-CD 1 to 3. We used this mAIS code, along with the other AIS codes, to calculate a separate NISSx value. If the resulting NISSx score falls into the highest S-scale cluster (see below), the case was included in the evaluation.[11]

##### AIS 9 Code: non-survivable Injury

Each AIS-ID coding was scanned for a non-survivable injury or an injury that, by description, was not survivable, e.g., ‘torso transsection,’ or ‘died of head injuries.’ All patients with a non-survivable AIS 9 coding were assigned a HARM 4 rating.

Note that this is a conservative estimate, i.e., a possible overestimation of the HARM-level, as some of the deceased may have been HARM 3, and a departure from the probabilistic AIS scale. It was solely used to recruit more AIS 9 cases for the evaluation.

#### 3.2.2. FCI

##### FCI-Code Aggregation

While the most naïve way of aggregating FCI-CD to the person level would be to use the maximum FCI value across all the persons’ injuries, this ‘winner takes all’ approach has been shown in the AIS-CD case, i.e., the MAIS, to be bested by using the three most severe injuries to calculate an injury severity. Therefore, we used the AIS-CD aggregation metric based on triples, as detailed above, and adapted it to the FCI codes.

FCI-CD aggregation was not possible in patients with even a single FCI 9 code because, unlike the AIS-CD, a reliable minimum FCI value could not be derived. However, to ensure the most severe outcomes were captured, a liberal aggregation approach was implemented for the person-level HARM assessment. If the injury aggregation process involved FCI 9 codes (which were temporarily treated as FCI 1 codes for this check) and the patient’s most severe injury outcome was still determined to be HARM 4 based on the overall result, an aggregated HARM value was successfully derived. For the remaining FCI 9 cases, only the AIS-CD was used in the final HARM determination.

### 3.3. Assignment of Triplets to HARM-Levels

#### 3.3.1. Assignment Method

While AIS-IDs are injury descriptors at the individual injury level, the HARM-levels describe injury severity at the person level. The chosen aggregation mirrors the New Injury Severity Score (NISS)[17] aggregation: the most severely rated injuries for each measure are grouped into AIS-CD and FCI-CD triples. These triples are assigned to the HARM-levels using a direct HARM-to-AIS/FCI triple match (see Table 2).

Note that the HARM-level is calculated independently for AIS-CD and FCI-CD, meaning it is assessed separately for mortality and long-term consequences. The resulting HARM-level assigned to a person is the most severe HARM-level from either AIS-CD or FCI-CD.

#### 3.3.2. Specific Assignment to HARM-Levels

There are four HARM severity levels in RAPEX, beginning with HARM 1 (minor) to HARM 4 (life-threatening). At the HARM 1 level, all injuries are easily treatable and do not necessarily need the attention of a medical doctor. Furthermore, the consequences of the injury are [almost] entirely reversible. This description aligns with S 0, S 0.5, and the respective FCI-CD triple.

A medical doctor must evaluate HARM 2-level (moderate) injuries, though hospitalization is usually not required. This description aligns with AIS-CD 2 injuries; however, patients with three or more AIS-CD 2 injuries typically require hospital care. Therefore, S 1 and S 1.5 align with HARM 2. The long-term impairment at HARM 2 should be ‘reversible within a certain period’, matching the corresponding FCI-CD triple.

At the HARM 4 level, the injuries start to be life-threatening, corresponding to AIS-CD 3 injuries, when combined with at least another moderate (AIS-CD 2) or more severe injury. Thus, the S 2.5 and S 3 injury severities are coded as HARM 4. Regarding long-term impairment, HARM 4 requires a ‘large negative effect’ that is ‘irreversible in several aspects,’ aligning with FCI-CD 3 (severe state) and FCI-CD 4 (worst possible state).

HARM 3 bridges the gap between HARM 2 and 4: S 2 and the corresponding FCI-CD triple.

## 4. Results

The mortality rate for different injury severities, stratified by AIS-ID levels, is shown in Figure 1.

In Table 3, the reduction in AIS-IDs across the aggregation levels is quantified for AIS Levels 1-3 and FCI Level 1. The AIS-CD 0s are a result of the purging of the mostly self-reported superficial skin injuries from the codebook. In the AIS-CD 9 columns, the minimum injury severity needed to sustain said injury is documented (see Method section).

**Table 3:** Number of codeable AIS-IDs for the different AIS-CD and FCI-CD levels. Note: The AIS-CD 0 are purged superficial skin injuries, downcoded from AIS-CD 1. The AIS-CD 9 column states the minimal AIS-CD required to sustain the described injury.

| | CD | | | | | | | 9 | | | $\Sigma$ |
| --- | --- | --- | --- | --- | --- | --- | --- | --- | --- | --- | --- |
|  | 0 | 1 | 2 | 3 | 4 | 5 | 6 | 1 | 2 | 3 |  |
| AIS 2015 |  | 455 | 721 | 449 | 176 | 127 | 35 | 22 | 5 | 16 | 2006 |
| AIS 2015 LVL3 | 108 | 313 | 507 | 249 | 91 | 56 | 19 | 24 | 6 | 15 | 1388 |
| AIS 2015 LVL2 | 28 | 142 | 227 | 97 | 28 | 17 | 8 | 24 | 6 | 14 | 591 |
| AIS 2015 LVL1 | 17 | 82 | 117 | 54 | 12 | 11 | 8 | 24 | 6 | 14 | 345 |
| FCI 2015 |  | 1361 | 194 | 124 | 135 | 150 |  |  | 42 |  | 2006 |
| FCI 2015 LVL1 |  | 247 | 40 | 36 | 53 | 35 |  |  | 42 |  | 453 |

As the practical part of our investigation focuses on the AIS-Level 1 and FCI-Level 1 aggregations, the distribution of AIS-CDs over FCI-CDs is shown in Table 4. Even at FCI-Level 1, over 70% (1430/2026) of the injuries have no or only very minor long-term consequences. Of the 1979 non-9 codes, 25.3% are identical between AIS and FCI. In 25.8% of cases, the FCI-CD is more severe than the AIS-CD, and in 48.9% the reverse is true.

**Table 4:** AIS-IDs: Correlation of AIS-CDs and FCI-CDs at Level 1. Note: 20 new AIS-IDs were introduced to better differentiate between AIS and FCI in Level 1 aggregation, increasing the number of AIS-IDs from 2006 to 2026.

| | | AIS 2015 Level 1 CD | | | | | | | 9 | | | $\Sigma$ |
| --- | --- | --- | --- | --- | --- | --- | --- | --- | --- | --- | --- | --- |
|  |  | 0 | 1 | 2 | 3 | 4 | 5 | 6 | 1 | 2 | 3 |  |
| FCI-CD Level 1 | 1 | 148 | 366 | 564 | 319 | 24 | 7 |  | 2 |  |  | 1430 |
|  | 2 | 1 | 52 | 88 | 43 |  |  |  |  |  |  | 184 |
|  | 3 | 2 | 35 | 49 | 21 |  |  |  |  |  |  | 107 |
|  | 4 | 1 | 19 | 41 | 52 | 6 | 2 |  |  |  |  | 121 |
|  | 5 |  | 2 | 20 | 86 | 3 | 19 | 9 |  |  |  | 139 |
|  | 9 |  |  |  |  |  |  |  | 25 | 6 | 14 | 45 |
| $\Sigma$ | | 152 | 474 | 762 | 521 | 33 | 28 | 9 | 27 | 6 | 14 | 2026 |

The application of the HARM calculation to the GIDAS dataset is detailed in Table 6. For lower HARM-levels, FCI-based long-term consequences appear to have a limited influence, as we are examining only injured persons, i.e., those with at least an AIS-CD 1 injury. Furthermore, injuries rated low on the AIS scale are either also low on the FCI scale (14%) or have a significantly higher FCI rating, thereby shifting the injury to HARM-levels 3 or 4.

At HARM 2, 15% of patients experience injuries with long-term consequences, even though in only 1% of these cases do the long-term consequences define the HARM severity. At HARM 4, AIS-CD is the driving force in 53% of cases, while FCI is the driving force in 16%. For the remaining 31% of cases, both AIS and FCI point to HARM 4.

As HARM severity increases, long-term consequences take on a more defining role.

Examples of injuries with a ‘low’ AIS-CD and a higher-rated FCI-CD are given in Table 5.

**Table 5:** Examples of Injuries with a low AIS-CD and a high FCI-CD rating. Note that HARM is an aggregated injury scale, so its value depends on other injuries. The HARM values given here are if no other or only further minor injuries are present.

| injury description | AIS codebook |  | Level 1 aggregation |  | HARM |  |
| --- | --- | --- | --- | --- | --- | --- |
|  | AIS-CD | FCI-CD | AIS-CD | FCI-CD | AIS | FCI |
| hypoglossal nerve (XII) laceration | 2 | 4 | 2 | 4 | 2/3 | 4 |
| brachial plexus, complete plexus injury | 3 | 5 | 2 | 5 | 2/3 | 4 |
| knee joint dislocation | 2 | 4 | 1 | 4 | 1 | 4 |
| knee joint avulsion patella | 2 | 5 | 1 | 5 | 1 | 4 |
| closed tibia fracture, complete articular | 2 | 4 | 2 | 4 | 2/3 | 4 |

**Table 6:** Decomposition of the HARM-levels for the GIDAS dataset. Which part of the HARM metric, lethality (AIS) or LTC (FCI), contributed what to overall HARM. *n:* number of cases in GIDAS; *p:* percentage within HARM-level; *dominant metric:* which metric was pushing the HARM-level upwards in percentage.

| HARM | HARM decomposition |  | n | p | dominant metric [%] |  |  |
| --- | --- | --- | --- | --- | --- | --- | --- |
|  | AIS part | FCI part |  |  | AIS-CD | none | FCI-CD |
| 1 | 1 | 1 | 88112 | 100 | 0 | 100 | 0 |
| 2 | 1 | 2 | 47 | 1 | 85 | 14 | 1 |
|  | 2 | 1 | 7196 | 85 |  |  |  |
|  | 2 | 2 | 1264 | 15 |  |  |  |
| 3 | 1 | 3 | 70 | 5 | 80 | 7 | 13 |
|  | 2 | 3 | 125 | 9 |  |  |  |
|  | 3 | 1 | 726 | 50 |  |  |  |
|  | 3 | 2 | 433 | 30 |  |  |  |
|  | 3 | 3 | 98 | 7 |  |  |  |
| 4 | 1 | 4 | 18 | 1 | 53 | 31 | 16 |
|  | 2 | 4 | 260 | 9 |  |  |  |
|  | 3 | 4 | 185 | 6 |  |  |  |
|  | 4 | 1 | 870 | 29 |  |  |  |
|  | 4 | 2 | 410 | 14 |  |  |  |
|  | 4 | 3 | 160 | 5 |  |  |  |
|  | 4 | 4 | 918 | 31 |  |  |  |
|  | 4 | 9 | 150 | 5 |  |  |  |
|  | 9 | 4 | 7 | 0 |  |  |  |

The special treatment of AIS-CD 9-coded injuries successfully increased the case count for the highest S-scale cluster (S 2.5 and S 3.0). Specifically, 191 individuals were assigned to HARM 4 among the 4,701 patients with at least one of the 5,270 AIS-CD 9 injuries in GIDAS. Since a comparable minimum value could not be derived for the FCI, only a liberal inclusion of FCI-CD 9 was possible, resulting in 76 of the 191 overlapping cases and only seven additional individuals in HARM 4. Consequently, the HARM 4 ratings exhibit a bias toward the survivability metric (AIS-CD) due to this differential treatment of ‘unknown’ severity codes.

## 5. Discussion

Aligning the level of medical detail required for AIS-ID coding with the medical diagnoses necessary for treatment improved the correlation between the resulting AIS-CDs and mortality, as AIS-ID coding relies on medical diagnostics used for treatment and diagnosis. However, there is a lack of medical diagnostics specific to AIS coding. Thus, a reduction in the level of detail results in an improved AIS code.

Building on the knowledge about the correlation between real-world medical diagnostics and mortality, we were able to define three levels of abstraction for the AIS code. Information on crash-related functional incapacitation, its diagnosis, which is primarily based on physical examination and medical imaging, and its long-term consequences is scarce. Thus, we only describe one level of AIS-ID aggregation for FCI evaluation.

It should be noted that the AIS-ID aggregation for AIS-CD and FCI-CD evaluation differs, even when comparing Level 1 AIS- and FCI-focused AIS-ID aggregations. It is basically the same data, i.e., AIS-IDs, but different clustering methods.

The aggregation to Level 1 alters the AIS-CDs and FCI-CDs. It is seen as a measure to better align injuries with medical reality: it is based on what can actually be extracted from the given medical documentation, i.e., its goal is to reduce up- and downcoding of injury severity in AIS-CD and FCI-CD.

We have shown how to post-hoc assign HARM values to AIS 2015-coded injuries, opening crash and accident databases for RAPEX evaluations. Furthermore, the HARM values, with their unique mix of mortality and long-term consequences, can be treated as a measure of medical injury severity and used for evaluating crash scenarios or crash configurations. Thus, it offers a more holistic approach than the current AIS-CD-only, mortality-driven view. An application could be HARM-driven ISO26262 S-parameters.[11]

## Supporting information

AIS 2025 LVL

## Data Availability

Authors do not have permission to share the data.

## 6. Limitations

While a strong correlation between AIS-CD and mortality has been demonstrated in numerous studies, including this one, a similar correlation has not yet been established for AIS 2015-based FCI-CD and long-term consequences. Although there are hints of the usefulness of the FCI, datasets that prove the correlation seem scarce.

Furthermore, the comparability of long-term consequences across body regions seems complicated: how can one compare diminished cognitive function with the inability to walk without painkillers?

## Notes

### Competing Interest Statement

The authors have declared no competing interest.

### Funding Statement

This study did not receive any funding

### Author Declarations

The data comes from the GIDAS project. https://www.gidas.org/ The German In-Depth Accident Study (GIDAS) dataset is strictly de-identified and anonymized before researchers are ever granted access to it. How GIDAS De-Identifies Data To comply with strict German data privacy laws and the European General Data Protection Regulation (GDPR), the GIDAS database anonymizes all cases immediately after data collection. Removal of Personal Details: Names, addresses, and exact dates of birth of the persons involved in the accident are never stored in the database. Removal of Unique Identifiers: To prevent anyone from linking a specific database entry to a real-world news story or police report, GIDAS does not store exact accident times, license plate numbers, or chassis numbers (VINs). Visual Censoring: In all photographic and video evidence collected at the scene, the faces of individuals, license plates, and unique vehicle features (like specific company logos or custom decals) are heavily blurred and made unrecognizable

